# An age-structured epidemiological model of the Belgian COVID-19 epidemic

**DOI:** 10.1101/2020.04.23.20077115

**Authors:** Koen Deforche

## Abstract

COVID-19 has prompted many countries to implement extensive social distancing to stop the rapid spread of the virus, in order to prevent over-loading health care systems. Yet, the main epidemic parameters of this virus are not well understood. In the absence of broad testing or serological surveillance, it is hard to evaluate or predict the impact of different strategies to exit implemented lock-down measures. An age-structured epidemiological model was developed, which distinguishes between the younger versus older population (e.g. < 65 and ≥ 65). Because the illness severity is markedly different for these two populations, such a separation is necessary when estimating the model based on death and hospitalization incidence data. The model was applied to data of the Belgian epidemic and used to predict how the epidemic would react to a relaxing of social distancing measures.

## 1 Introduction

In previous analyses of the COVID-19 epidemic in several Asian and European countries, epidemiological parameters have been estimated by matching the observed growth of positive confirmed tests and/or deaths to standard epidemiological models [Prem et al., 2020]. One such often used class of models are compartment models, which use differential equations to describe in a deterministic way how individuals move between different compartments, each of which indicates a certain disease state. Standard compartment models are SIR or SEIR models, which use up to four compartments: Susceptible (“S”), Exposed (“E”), Infectious (“I”), and Removed (“R”). If however the transmission of the virus is different in the younger and older population, and also the incidence and mortality differs between these age groups, the main assumptions for estimating such models from case data are violated and conclusions drawn from such estimates may be unfounded.

By dividing the population in age-groups, there are multiple benefits:

- The model may assume different transmission dynamics within and between each age-group. For example, such a model may assume that transmission of the virus in the older group(s) is lower than transmission in the younger groups, for example because of different social interaction patterns.
- The model may match observed case reports and death incidence assuming an infection fatality rate that is suitable for the age-group.
- The model may be used to evaluate changes in social distancing policies that impact different age-groups differently.

In this analysis, we used an epidemiological model which employs SEIR models within each age-group, and allowing infectious individuals in one age-group infect susceptible individuals in another age-group. We add to a standard SEIR model the process in which infections give rise to hospitalizations and reported deaths. Rather than adding more compartments to the model, we model the number of hospitalizations as an independent process proportional to the incidence of an infection (when an individual moves from the “S” to “E” compartment), with a time delay. Likewise, reported deaths are modeled based on the removal of the infected individual (to the “R” compartment). The rate at which infections lead to deaths (the Infection Fatality Rate, IFR) may be different in each age-group.

In an SEIR model, changes in basic reproduction rate, because of changes in population behavior (such as the extreme social distancing measures that are being implemented) can be directly modeled by adding more parameters that describe changes in the basic reproduction number *R* within age-groups and between age-groups. We considered the Belgian epidemic with two age-groups: a “younger group” *y* (< 65) and an “older group” *o* (≥ 65). This division coincides more or less with a division of the active and retired population, in which one may expect different patterns of social interactions, leading to different transmission dynamics. Deaths reported in elderly care facilities (and which were in many cases not confirmed to be caused by COVID-19), were removed from the analysis, as the transmission into and within elderly care facilities is most likely quite different from elderly people living at home. Although about 100000 people older than > 65 are living in Belgian care centers (estimated by extrapolating percentage statistics from Flanders to the entire country), over 25 times as many do not. The model structure is shown in Figure 1.

**Figure 1:**
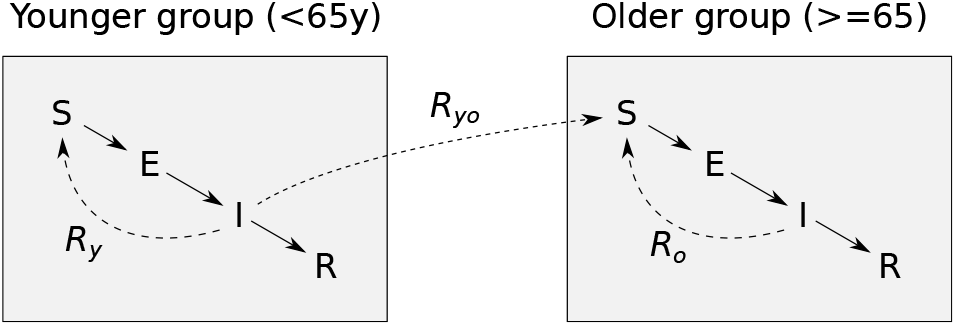
Model structure. Within each age group, individuals are considered to be in one of four states: susceptible (“S”), infected but latent (“E”), infectious (“I”) or recovered/death (“R”). Within each age group, infectious people can infect susceptible people, modulated by reproduction number *R*_*y*_ for the younger group, and *R*_*o*_ for the older group. The younger infectious people can also infect older people at rate *R*_*yo*_.

The Belgian government announced far-reaching social distancing on March 13th, effective from March 14th midnight, and added further measures on March 20th. This is modeled as shown in Figure 2.

**Figure 2:**
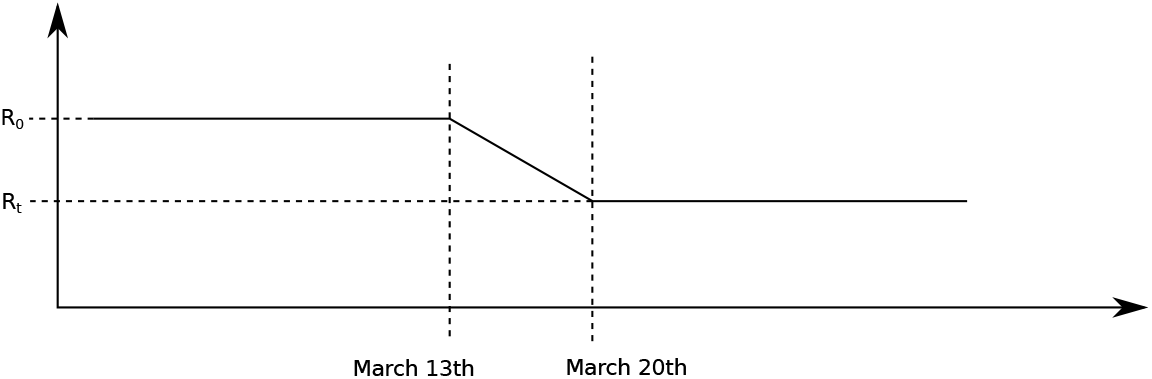
Piece-wise linear time profile of *R*_*y*_, *R*_*o*_ and *R*_*yo*_. The initial value *R*_*o*_ changes over a period of 7 days to a new value *R*_*t*_.

Parameters of this model were estimated using the open data provided by the Belgian government institute Sciensano. The model was fitted using a Bayesian framework which yields posterior credibility intervals for the key parameters, but also results in a functional model to make predictions for the future, taking into account all uncertainty of the underlying parameters.

### Below we list the main assumptions being made

- It is not possible to estimate the Infection Fatality Rate from the data (currently), and therefore, the estimate from Verity et al. [2020] wer used for each age group (taking into account the Belgian demography). In case the IFR is in fact lower (overall, or in one of the age groups), then the epidemic may take a whole different (less pessimistic) course than the one predicted by the current results.
- The control measures that were announced by the government have an impact on basic reproduction number *R*_0_ following a piece-wise linear model shown in Figure 2: a constant *R*_0_ before March 13th, a constant *R*_*t*_ after March 20th, and a linear interpolation between these two during the week between March 13th and March 20th.
- In each age-group, observed deaths are a fraction (the Infection Fatality Rate, IFR) of the “R” compartment, with a time delay *𝒩* (*µ*_*d*_, *σ*_*d*_).
- In each age-group, observed hospitalizations are a fraction (the Hospitalization Rate *H*) of the people that became infected (transition from “S” to “E” compartments), with a time delay *𝒩* (*µ*_*h*_, *σ*_*h*_).

## 2 Methods

### 2.1 Model

The following differential equations describe the dynamics of individuals in each of the compartments of Figure 1.

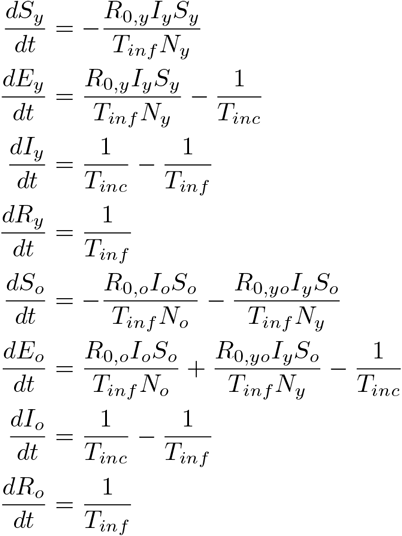

These equations use the following parameters:

- *R*_0_, the basic reproduction number: the average number of new infections caused by each given person provided all people are susceptible (thus actual reproduction number decreases linearly as the number of susceptible people decreases). The value of *R*_0_ can vary over time, for example because of measures taken to contain the virus.
- *T*_*inf*_, the average period that a person is infectious (stays in the “I” compartment)
- *T*_*inc*_, the average time to become infectious. This may not be different from the clinical incubation time, which is the time to start showing symptoms.

Table 1 lists all parameters and their values (either a constant, or a prior distribution for parameters that are estimated).

**Table 1:**
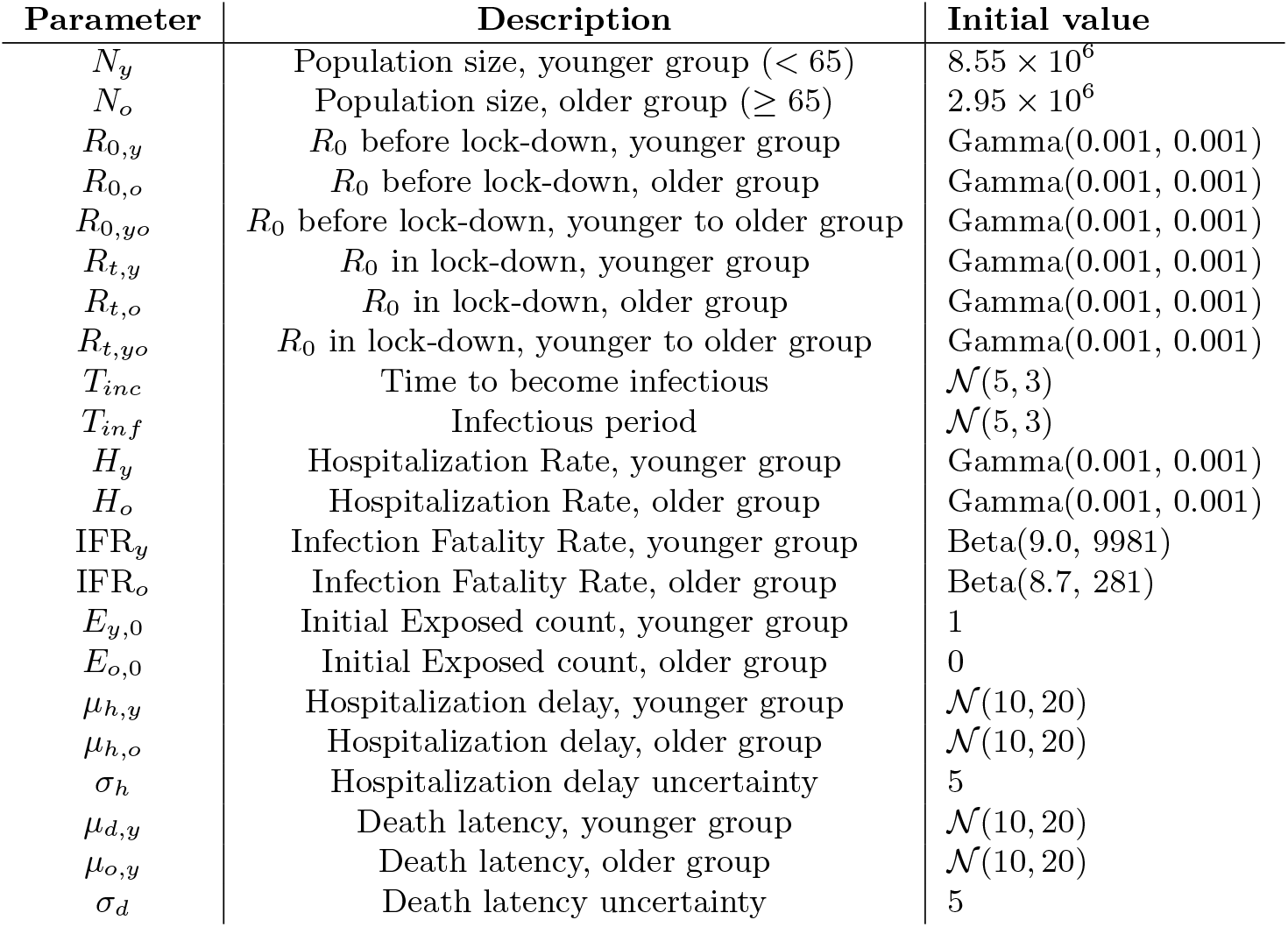
Parameters of model

| Parameter | Description | Initial value |
| --- | --- | --- |
| $N_y$ | Population size, younger group ($< 65$) | $8.55 \times 10^6$ |
| $N_o$ | Population size, older group ($\geq 65$) | $2.95 \times 10^6$ |
| $R_{0,y}$ | $R_0$ before lock-down, younger group | Gamma(0.001, 0.001) |
| $R_{0,o}$ | $R_0$ before lock-down, older group | Gamma(0.001, 0.001) |
| $R_{0,yo}$ | $R_0$ before lock-down, younger to older group | Gamma(0.001, 0.001) |
| $R_{t,y}$ | $R_0$ in lock-down, younger group | Gamma(0.001, 0.001) |
| $R_{t,o}$ | $R_0$ in lock-down, older group | Gamma(0.001, 0.001) |
| $R_{t,yo}$ | $R_0$ in lock-down, younger to older group | Gamma(0.001, 0.001) |
| $T_{inc}$ | Time to become infectious | $\mathcal{N}(5, 3)$ |
| $T_{inf}$ | Infectious period | $\mathcal{N}(5, 3)$ |
| $H_y$ | Hospitalization Rate, younger group | Gamma(0.001, 0.001) |
| $H_o$ | Hospitalization Rate, older group | Gamma(0.001, 0.001) |
| $IFR_y$ | Infection Fatality Rate, younger group | Beta(9.0, 9981) |
| $IFR_o$ | Infection Fatality Rate, older group | Beta(8.7, 281) |
| $E_{y,0}$ | Initial Exposed count, younger group | 1 |
| $E_{o,0}$ | Initial Exposed count, older group | 0 |
| $\mu_{h,y}$ | Hospitalization delay, younger group | $\mathcal{N}(10, 20)$ |
| $\mu_{h,o}$ | Hospitalization delay, older group | $\mathcal{N}(10, 20)$ |
| $\sigma_h$ | Hospitalization delay uncertainty | 5 |
| $\mu_{d,y}$ | Death latency, younger group | $\mathcal{N}(10, 20)$ |
| $\mu_{d,o}$ | Death latency, older group | $\mathcal{N}(10, 20)$ |
| $\sigma_d$ | Death latency uncertainty | 5 |

The SEIR model equations were calculated using a finite difference approximation with time steps of one day. The cumulative values for hospitalizations and deaths were calculated by convolving the normal density function *𝒩* (*µ*_*h*_, *σ*_*h*_) and 𝒩 (*µ*_*d*_, *σ*_*d*_) over the obtained estimates of *N*−*S* and *R* respectively, within each age group.

To fit the model against the data, a likelihood calculation is done for each incidence data point (number of hospitalizations per day, or number of deaths per day, within each age group), using a negative binomial distribution with *µ* equal to the expected count, with a dispersion parameter *r* that reflects the observed clustering in the data. Clustering may be present because some hospitals are not reporting each day (typically seen for weekends). The dispersion parameters *r* for the negative binomial distributions were estimated based on the observed variance in the data. This was done by observing the model prediction of a maximum likely model *µ* with the observed data *s*, and setting the dispersion parameter to a value about 10% lower than the *r* size value for which the following relation holds, to obtain a measured variance that matches the expected variance

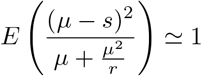

This resulted in a dispersion parameter value *r*_*h*_ = 35 for evaluating hospitalization incidence, and *r*_*h*_ = 90 for evaluating death incidence.

The posterior distribution of the model parameters was estimated using adaptive MCMC. Analyses, convergence diagnostics and inferences were run in R [Li et al., 2020, Scheidegger and and, 2018, Flegal et al., 2020, Makowski et al., 2019].

The R scripts are available at https://github.com/kdeforche/epi-mcmc.

### 2.2 Data

Data on the incidence of deaths and hospitalizations were based on data made available by Sciensano. Data up to April 19 for hospitalizations, and April 17 for deaths was used.

For hospitalizations, the published data sets do not provide a break down in age groups. In the epidemiological report of April 16, the age breakdown of hospitalized people is shown in two charts (chapter 2.5.1.2). Based on these charts, a breakdown of hospitalizations in the two age groups was estimated, as shown in Figure 3. A simple polynomial model was fitted to distribute the hospitalization data accordingly into the two age-groups.

**Figure 3:**
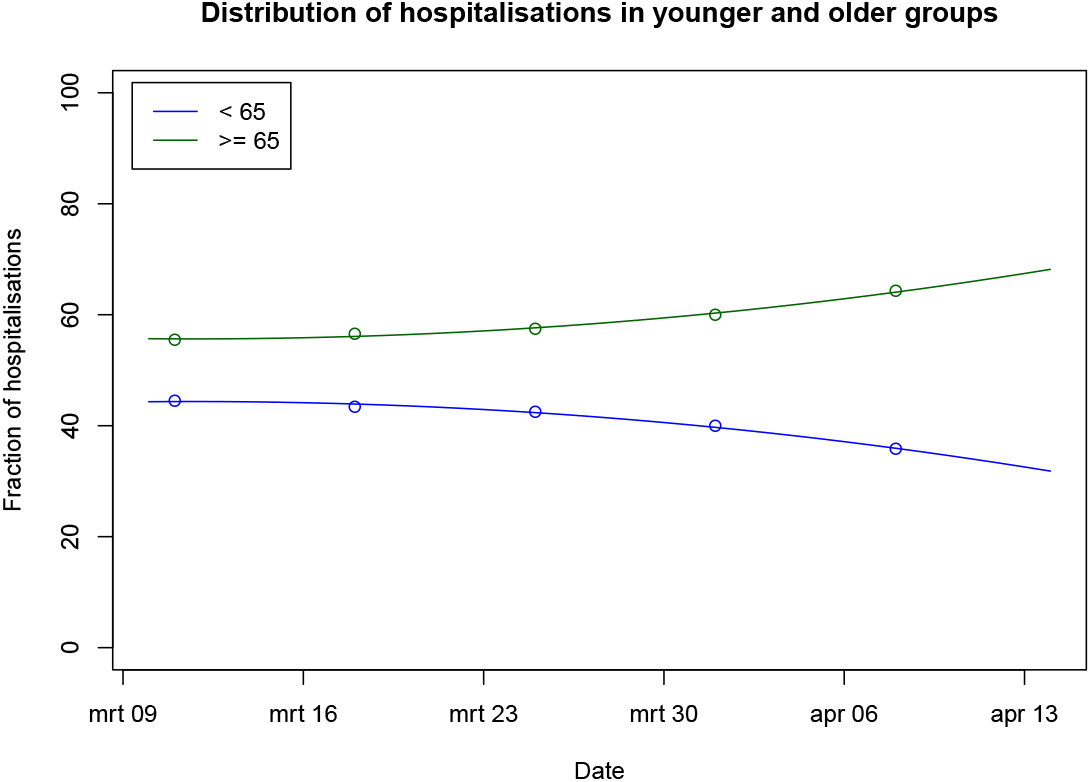
Age group distribution of hospitalizations.

For deaths, the published data sets includes a breakdown in age groups. To exclude deaths reported in elderly care centers, the data from the charts in the epidemiological report (chapter 2.7.1) was used to remove deaths reported in elderly care centers (which all belong to the older age group).

The prior distributions for IFR_*y*_ and IFR_*o*_ were calculated based on the age breakdown of IFR in Verity et al. [2020], applied to the age-breakdown of the two age groups in the Belgian population (excluding elderly care centers).

## 3 Results

Table 2 summarizes the posterior estimates for all parameters, and the values for *R*_0_ are illustrated in Figure 4.

**Table 2:** Parameter estimates with credible intervals (cri)

| Parameter | Median | 50% cri | 95% cri |
| --- | --- | --- | --- |
| $R_{0,y}$ | 4.1 | 3.7 – 4.6 | 3.1 – 5.7 |
| $R_{0,o}$ | 1.7 | 0.7 – 3.2 | 0.1 – 5.3 |
| $R_{0,yo}$ | 0.8 | 0.3 – 1.7 | 0.0 – 6.3 |
| $R_{t,y}$ | 0.8 | 0.7 – 0.8 | 0.7 – 0.9 |
| $R_{t,o}$ | 0.3 | 0.1 – 0.5 | 0.0 – 0.8 |
| $R_{t,yo}$ | 0.5 | 0.3 – 0.8 | 0.0 – 2.0 |
| $T_{inc}$ | 5.9 days | 5.0 – 6.7 | 2.1 – 8.1 |
| $T_{inf}$ | 1.2 days | 1.1 – 1.5 | 1.1 – 2.5 |
| $H_y$ | 0.8% | 0.7 – 1.1 | 0.4 – 1.6 |
| $H_o$ | 5% | 3 – 7 | 2 – 11 |
| $IFR_y$ | 0.07% | 0.05 – 0.09 | 0.03 – 0.14 |
| $IFR_o$ | 1.8% | 1.3 – 2.4 | 0.6 – 3.8 |
| $\mu_{h,y}$ | 10 days | 9 – 11 | 9 – 11 |
| $\mu_{h,o}$ | 9 days | 7 – 10 | 4 – 11 |
| $\mu_{d,y}$ | 12 days | 12 – 13 | 11 – 14 |
| $\mu_{d,o}$ | 8 days | 7 – 10 | 1 – 10 |

**Figure 4:**
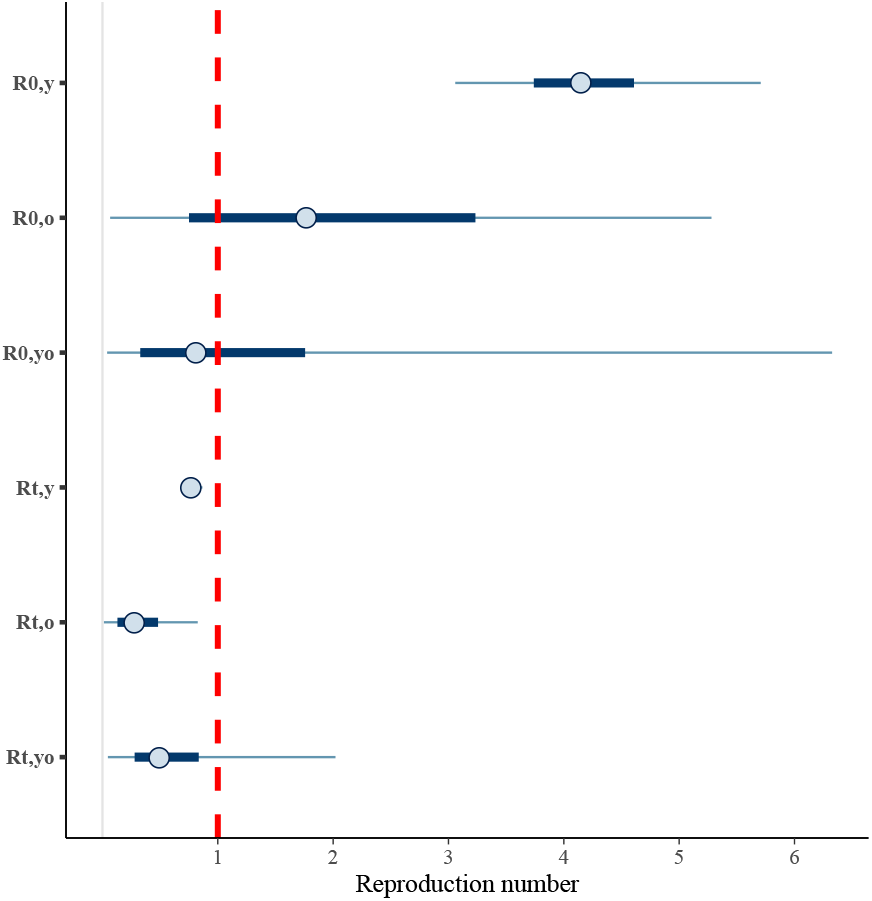
Posterior estimates of reproduction numbers *R*_0_ (before lock-down) and *R*_*t*_ (in lock-down)

The estimate of the current state of the epidemic, and predicted evolution in case current measures are being continued, are shown in Figure 5.

**Figure 5:**
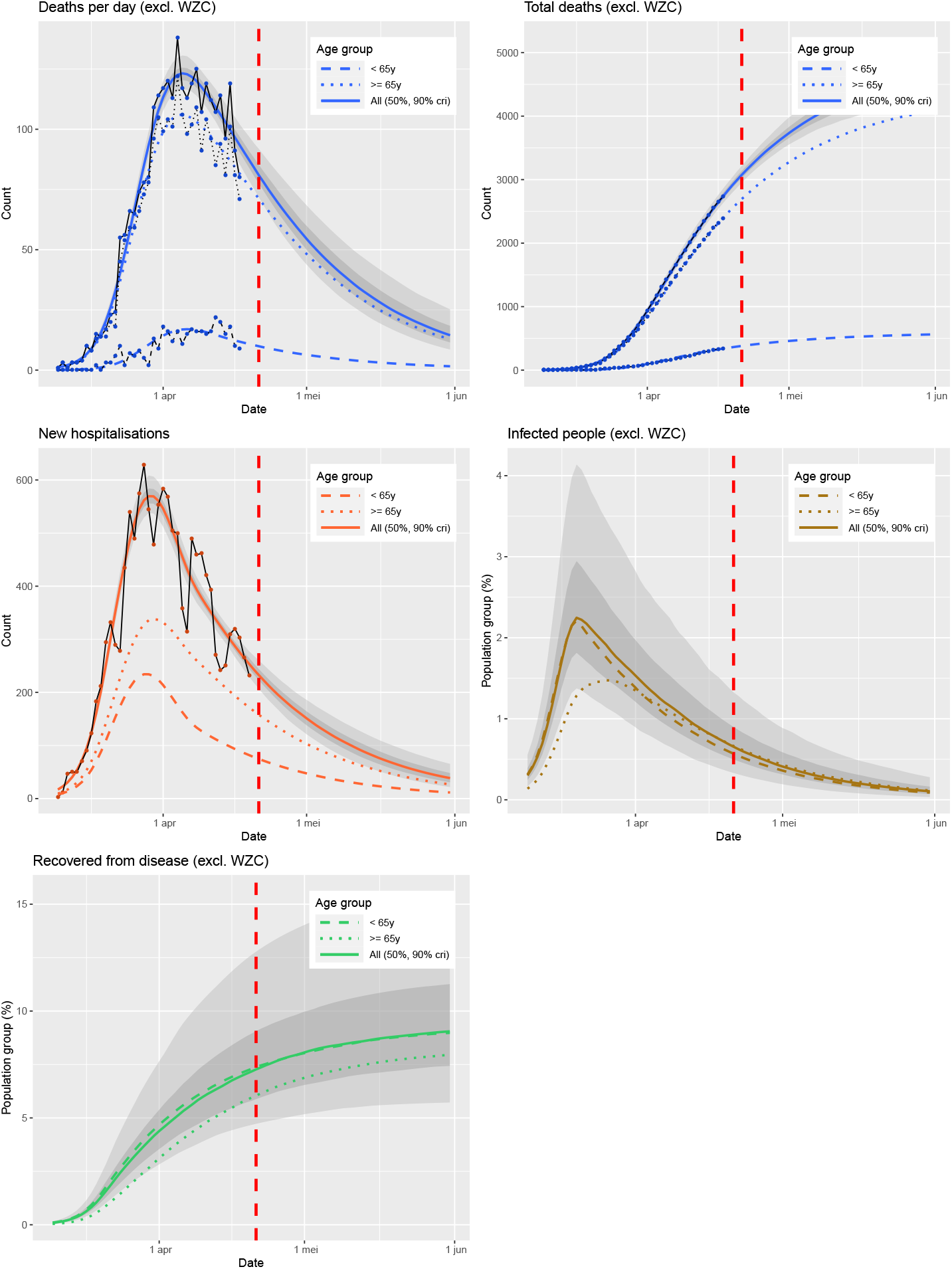
Estimated current state of the Belgian epidemic, and short-term evolution provided the current measures are being continued. The red line marks April 21.

Figures 6 and 7 show a predicted evolution of the epidemic when the measures for the younger group would be relaxed on May 4, by respectively 15, and 20% compared to the current level, while keeping the same social distancing withing the older group, and between the younger and the older group. Likewise, figure 8 shows the effect if the measures would be entirely lifted within the younger group.

**Figure 6:**
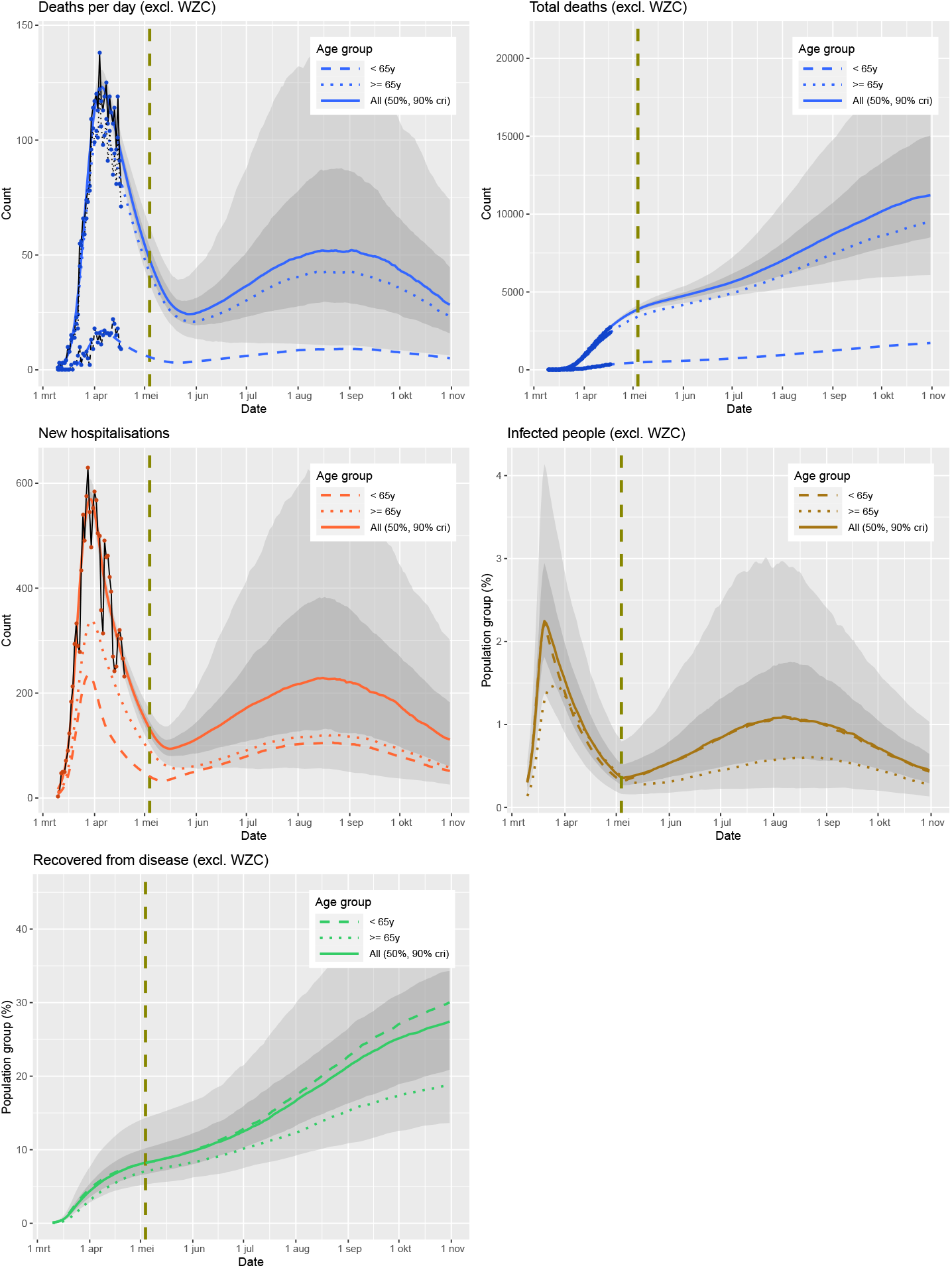
Estimated evolution of the Belgian epidemic, under a scenario where the current measures are being relaxed for the younger group by 15%, but kept the same within the older group, and between the two age-groups. The line marks May 4, the simulated start date for relaxing the measures.

**Figure 7:**
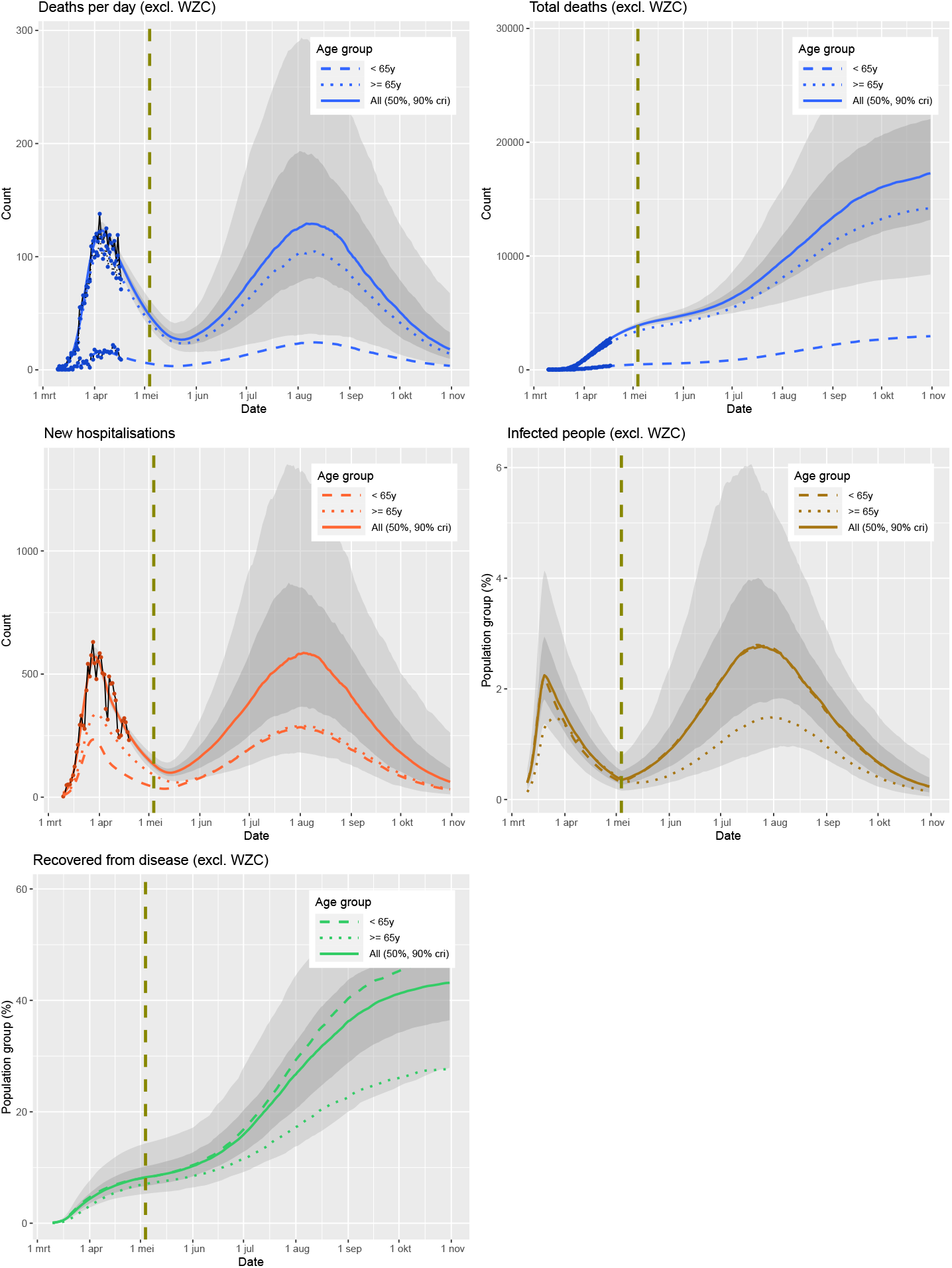
Estimated evolution of the Belgian epidemic, under a scenario where the current measures are being relaxed for the younger group by 20%, but kept the same within the older group, and between the two age-groups. The line marks May 4, the simulated start date for relaxing the measures.

**Figure 8:**
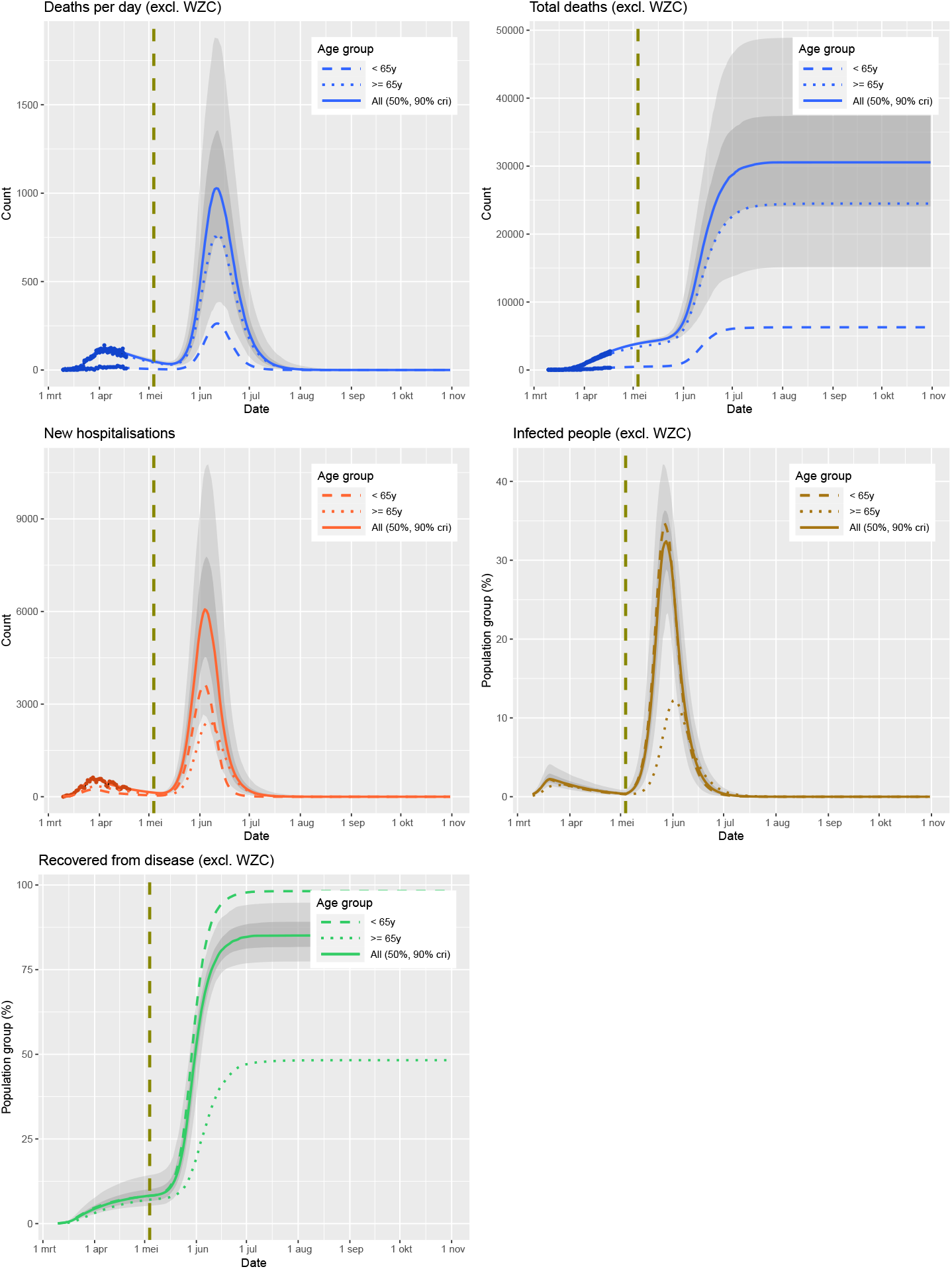
Estimated evolution of the Belgian epidemic, under a scenario where the current measures are being lifted for the younger group, but kept the same within the older group, and between the two age-groups. The line marks May 4, the simulated start date for relaxing the measures.

## 4 Discussion

A model was developed for modeling a COVID-19 epidemic, based on incidence data of hospitalizations and deaths. To account for the difference in clinical outcome of the infection in younger versus older age-groups, epidemiological parameters were estimated for the two age-groups separately. The model considers that younger people can infect older people, but infections of the younger group by the older group was omitted to limit the number of model parameters to be estimated, and as it was assumed that there would be a higher prevalence infections in the younger age group which is assumed to have a higher number of social interactions, and to limit the number of model parameters.

When applied to the Belgian epidemic, the model estimates that before lockdown, the virus had an *R*_0_ value of 4.1 (95% cri 3.1 – 5.7) in the younger age group, compared to 1.7 (0.1 – 5.3) in the older age group, and 0.8 (0.3 – 1.7) from the younger age group to the older age group. The large uncertainty on these parameter estimates is likely because only three days of hospitalization and death incidence data are available before lockdown, and thus all later data is confounded by the impact of lockdown. Because of the higher *R*_0_ value in the younger age group, it is estimated that the virus circulated more within the younger age group than the older age group, before lockdown, see Figure 5 (Infected people). Thus, social interaction within and between the older age group is by its own able to prevent to a large extent the rapid spread of the virus. The lockdown measures brought the basic reproduction number in the younger age group down to *R*_*t*_ = 0.8 (0.7 – 0.9), in the older age group to 0.3 (0.1 – 0.8) and from the younger to the older age group to 0.5 (0.0 – 2.0). By April 1, the prevalence of the virus is estimated to be similar in the two age groups, and declining. The measures thus brought the epidemic under control and the pressure on the health care system can be expected to decline as long as the lockdown measures are adhered to.

By accounting for the different age groups separately, scenario’s can be simulated for changing the measures where the social distancing behavior of the two different age groups is impacted differently. Examples of such simulations are shown in Figures 6, 7, and 8. It shows that a slight relaxing of the social distancing rules within the younger age group is feasible (in the sense that the health-care system is likelly not to be overrun) at the beginning of May. The evolution of the epidemic is predicted to have the majority of deaths in the older age group, even though the percentage of infections is predicted to be higher in the younger age group. The epidemic would reach a new peak in the summer. Herd immunity (even if limited), in combination with the social distancing, would then level off the epidemic. In such a scenario there thus would not need to be a new lockdown.

Deaths that occur within elderly care centers were excluded from this study. Nevertheless, hospitalization of residents of elderly care centers are a significant part of hospitalizations, and thus the epidemic within the care centers influences this analysis and results. The inferred results cannot be interpreted as applying solely to elderly people living outside elderly care centers. It is plausible that the isolation of elderly people outside ederly care centers is therefore underestimated, which would be beneficial to scenarios where social distancing is being relaxed within the younger age group but maintained (or enhanced) towards and within the older age group.

## 5 Conclusion

An epidemiological model for COVID-19 was developed which considers the epidemic within the younger age group and older age group separately. The model provides insight in the different evolution of the epidemic in these two age groups, and the interaction between the two age groups, and can be used to evaluate the impact of changes in social distancing measures that treat the two age groups differently.

## Data Availability

All data has been made openly available by the Belgian governement (Sciensano).

https://github.com/kdeforche/epi-mcmc

## 6 Acknowledgments

We are very grateful for the public sharing of data on the Belgian epidemic, organized by Sciensano, and possible through the daily efforts in Belgian hospitals and care centers.

